# Viewing Direct-to-Consumer Genetic Test Results for Depression Risk Is Psychologically Well Tolerated: Evidence from a Longitudinal Equivalence Study

**DOI:** 10.1101/2025.07.21.25330778

**Authors:** Rebecca M. K. Berns, Devika Dhamija, Daniella Coker, Chelsea L. Robertson, Jingran Wen, 23andMe Research Team, R. Ryanne Wu, Michael V. Holmes, Noura S. Abul-Husn

**Affiliations:** 23andMe, Inc., San Francisco, CA; Duke University School of Medicine, Durham, NC; Icahn School of Medicine at Mount Sinai, New York, NY

## Abstract

**Background:** Depression is a frequent focus of interest in genetic testing. Despite growing availability of polygenic risk scores (PRS) for depression, little is known about the psychological impact of receiving them in real-world settings. To quantify the impact of receiving an at-risk depression PRS result on depression and anxiety symptoms, we conducted a longitudinal, prospective cohort study of 23andMe, Inc. research participants.

**Methods:** Surveys were conducted between October 19, 2022 and October 9, 2023. Eligible participants were U.S. residents ≥ 18 years old who completed two surveys assessing depression and anxiety symptoms and had an at-risk PRS result for depression (odds ratio ≥ 1.5). We compared individuals who viewed their result to individuals who did not. Primary outcomes were changes in depression (Patient Health Questionnaire-8) and anxiety (Depression Anxiety Stress Scale-21) symptom scores relative to baseline. We fitted linear regressions to model each outcome, adjusting for age, sex, genetic ancestry, income, prior depression and anxiety, and baseline scores. Using an equivalence testing framework, the smallest effect size of interest was defined as Cohen’s d = ±0.5.

**Findings:** We analyzed data from 917 participants, including 361 who viewed the depression PRS and 556 who did not. Score changes from baseline to follow-up were statistically equivalent for individuals who viewed PRS results and those who did not. The adjusted between-group differences in score changes were −0.17 points for depression (90% CI, −0.59–0.24, two one-side tests p < 0.001) and −0.092 points for anxiety (90% CI, −0.35–0.17, two one-side tests p < 0.001), both equivalent within the predefined margin. Results were consistent in substrata defined by presence or absence of prior depression or anxiety.

**Interpretation:** Among genetically at-risk individuals, exposure to a depression PRS result was well-tolerated in a real-world setting.

**Funding:** 23andMe, Inc.

## Introduction

For most individuals, genetic risk for psychiatric conditions is not conferred by a single high-impact genetic variant but rather the cumulative impact of many genetic variants that individually have small effects on risk.(1) Polygenic risk scores (PRS) estimate the cumulative risk conferred by common variants, and can predict risk for anxiety, depression, and other psychiatric conditions.(2,3) Such PRS are becoming increasingly available in clinical, research, and consumer settings.(4,5) However, little is known about the psychological impact of receiving PRS results related to psychiatric conditions.(6–8)

Past studies, largely focusing on risk for cardiovascular disease and common cancers, have not demonstrated psychological harms of receiving PRS results.(6,9–12) This is consistent with earlier literature on genetic testing for monogenic conditions that found minimal and short-lived anxiety associated with delivery of at-risk results.(13) Psychiatric genetic testing warrants particular attention because of the sensitive nature of this information.(14) Individuals with current or past psychiatric conditions, or with genetic vulnerability to such conditions, may be particularly likely to experience negative psychological effects of receiving risk information(15) such as symptom exacerbations, while also being more motivated to seek genetic explanations for their personal and/or family history.(7,14,16–20) Few studies have examined the psychological impact of psychiatric genetic testing to date,(20–24) and only a handful of these were real-world studies.(20,24) A retrospective study found low levels of negative emotions in response to receiving a range of PRS that included psychiatric and other conditions, although a minority of participants reported high levels of negative emotions, with a lower understanding of PRS correlating with higher levels of negative emotions.(24) In a separate qualitative study of 18 individuals with bipolar disorder who could choose whether or not to receive a bipolar disorder PRS, while some individuals reported initial, transient distress, all individuals who chose to receive the PRS described the experience as a positive one that enhanced their understanding of the condition and reduced feelings of self-blame.(20) Together, these findings highlight the need for more prospective, real-world studies to understand how individuals respond to receiving psychiatric polygenic risk information.

We conducted a prospective cohort study—the Depression PRS Report Effect on Symptoms Study (DPRESS)—using longitudinal survey data to evaluate whether receiving a direct-to-consumer (DTC) genetic test report indicating elevated polygenic risk for depression is associated with changes in depression or anxiety symptoms. Our aim was to assess the psychological impact of receiving this information in a real-world setting, particularly among individuals identified as being at elevated genetic risk.

## Methods

### Participant recruitment

We recruited study participants from all genotyped 23andMe customers who opted to participate in research with 23andMe, Inc. All research participants were ≥ 18 years old, U.S. residents, provided informed consent and volunteered to participate in the research online, under a protocol approved by the external AAHRPP-accredited institutional review board Salus IRB (ethics approval number 10044; https://www.versiticlinicaltrials.org/salusirb). DNA extraction and genotyping were performed on saliva samples by Clinical Laboratory Improvement Amendments-certified and College of American Pathologists-accredited clinical laboratories of Laboratory Corporation of America. Samples were genotyped on custom Illumina genotyping arrays, as previously described(25).

### Eligibility criteria

Participants were eligible for the study if they resided in the US, were at least 18 years old and had elevated genetic risk for depression, as defined by an odds ratio ≥ 1.5 for a PRS predicting the likelihood of experiencing depression. Additionally, participants had to have completed a depression and anxiety survey in the six months prior to the exposure window (between October 19, 2022 and April 18, 2023, inclusive), and a follow-up instance of the same survey in the six months following the exposure window (between May 10, 2023 and October 9, 2023, inclusive).

### Depression PRS report

The depression PRS model was constructed as previously described,(26,27) using self-reported responses to previously deployed surveys that asked whether users have ever been diagnosed with, or treated for, depression. The PRS was constructed from a genome-wide association study (GWAS) using a separate cohort from the present study. The PRS model included 8,403 genetic variants, age, and self-reported sex, and was calibrated separately for individuals in East/Southeast Asian, European, Hispanic/Latino, Northern African/Central & Western Asian, South Asian, or Sub-Saharan African/African American genetic ancestry categories. We excluded participants whose PRS score may have changed due to methodological updates occurring during the study window.

Depression PRS results are delivered and presented within a standard electronic report (**eAppendix 1**) available through an on-line user account. The Depression PRS report, along with PRS reports on a wide range of other health conditions, are accessible to subscription members. The report provides a quantitative estimate of the user’s likelihood of being diagnosed with depression by age 30, a qualitative description of this risk that describes an odds ratio ≥ 1.5 as “increased likelihood” and odds ratios < 1.5 as “typical likelihood”, along with textual explanation and graphic illustration of these risk estimates. The report also includes information on the multifactorial nature of depression, explanation of non-genetic risk factors, possible ways to lower risk and manage symptoms, background on the condition, and a summary of scientific methods used to produce the result.

### Study arm definitions

At the time of analysis, 866,155 of the approximately 11 million 23andMe Research participants met inclusion criteria (**Figure 1**). Two study arms were defined. The exposed study arm consisted of participants who had access to the Depression PRS report based on service tier membership, who opted in to receive health reports, and who had opened and viewed the Depression report within three weeks of the report being made available within their 23andMe online account (the exposure window, April 19, 2023 – May 9, 2023). The unexposed study arm consisted of individuals who did not have access to the Depression PRS report, and thus could not have viewed their results.

**Figure 1.**
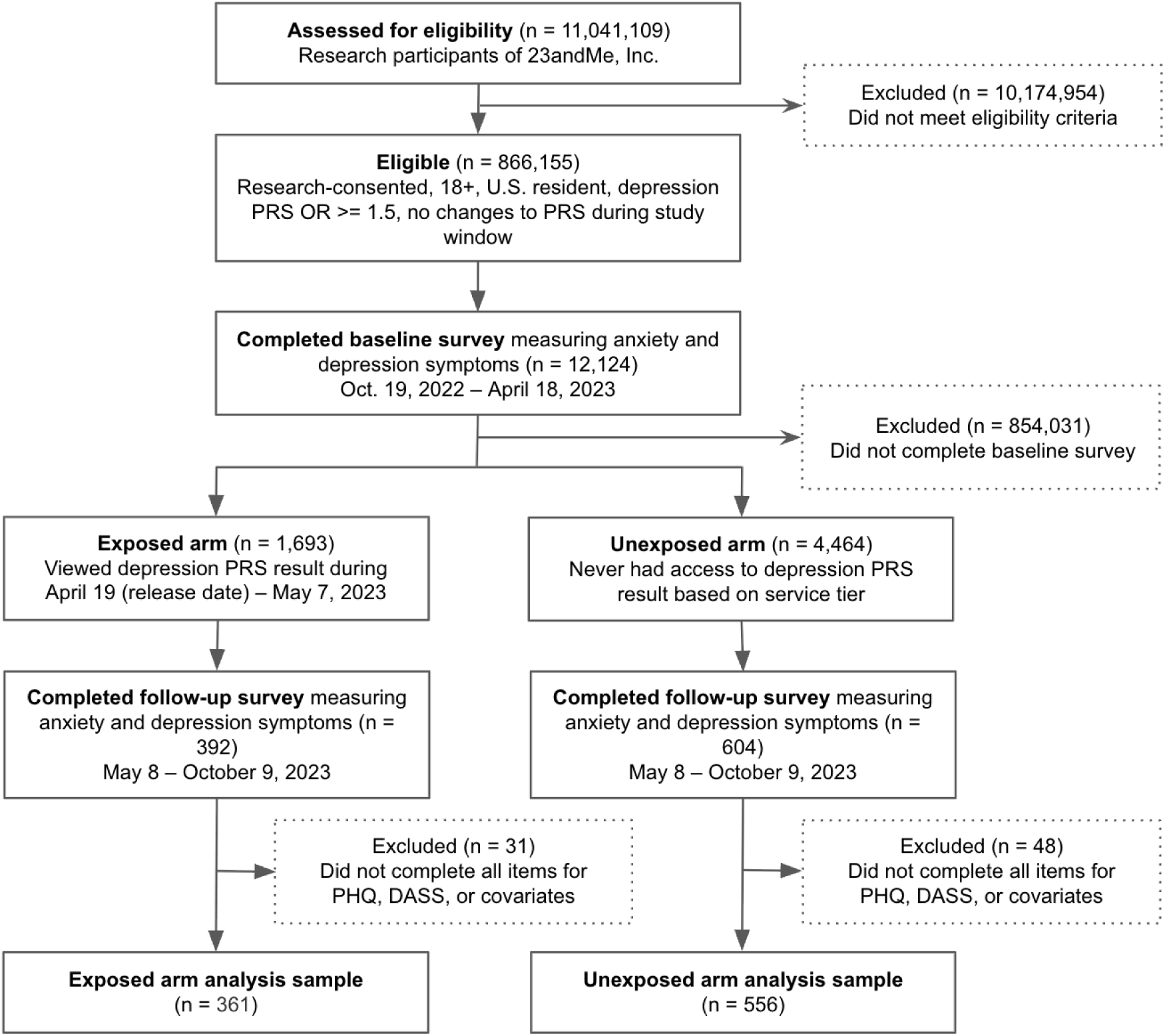
Flow of participants through the Depression PRS Report Effect on Symptoms Study (DPRESS), a longitudinal cohort study of 23andMe, Inc. research participants. Study flow diagram showing eligibility assessment, exclusions, and allocation to exposed (viewed a depression PRS result indicating increased likelihood of depression) and unexposed study arms. Participants completed symptom surveys between October 19, 2022 and October 9, 2023. The final analytic sample includes individuals with an at-risk PRS (OR ≥ 1.5) who completed baseline and follow-up symptom assessments.

**Figure 2.**
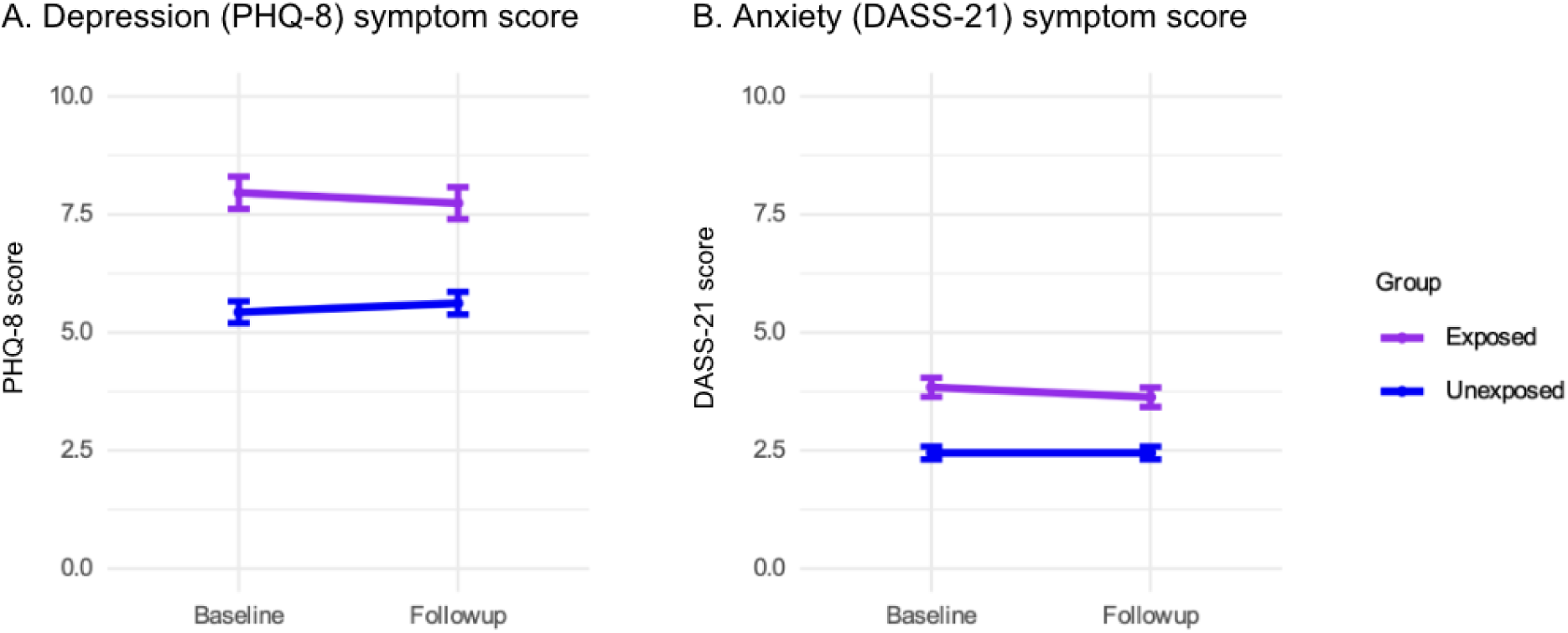
Depression and anxiety symptom scores from baseline to follow-up for exposed and unexposed study arms. A, Depression symptoms measured using the 8-item Patient Health Questionnaire (PHQ-8). B, Anxiety symptoms measured using the Depression Anxiety Stress Scales (DASS-21) anxiety subscale. Participants who viewed an at-risk depression PRS report (exposed) are compared with those who did not view the report (unexposed). Values shown are unadjusted (crude). Error bars indicate standard errors.

### Survey instrument

The study survey, a screening instrument assessing depression and anxiety symptoms, was made available to a subset of research participants in the Research tab of their online 23andMe accounts on a longitudinal basis. The survey consisted of 6 items from the anxiety subscale of the DASS-21 anxiety screening survey, and the PHQ-8 depression screening survey (**eAppendix 2**).(28,29) Both measures are widely used, well-validated self-report instruments with strong psychometric properties in general population and clinical samples, have been translated into a wide variety of languages, and are freely available in the public domain. The DASS-21 is a 21-item self-report questionnaire with three 7-item subscales measuring depression, anxiety, and stress severity. Commonly used score cut-offs for the anxiety subscale classify anxiety as normal (0–3), mild (4–5), moderate (6–7), severe (8–9), and extremely severe (≥10). The PHQ-8 is an 8-item self-report questionnaire measuring depression severity that is derived from the PHQ-9 by excluding the item on suicidal ideation. Commonly used score cut-offs classify depression as mild (5–9), moderate (10–14), moderately severe (15–19), or severe (20–24).

Individuals who completed one instance of the survey were subsequently emailed with invitations to complete up to three follow-up instances of the survey, at approximately six-month intervals. If participants completed more than one survey instance in either of these windows, the survey instance closer in time to the exposure was used.

Additional participant data collected via previously deployed surveys included age and median income at the level of zip code tabulation area (ZCTA, a proxy of socioeconomic status). Genetic ancestry groups were defined using 23andMe’s ancestry composition algorithm.(30)

### Statistical analyses

There were two primary outcome measures: the change in anxiety symptom scores from baseline to follow-up, and the change in depression symptom scores from baseline to follow-up. Each of the primary outcome measures was calculated for each study participant by subtracting the baseline value of each score from the follow-up value. We conducted a covariate-adjusted equivalence analysis to compare changes in depressive and anxiety symptoms between at-risk individuals who viewed their depression PRS results and at-risk individuals who did not. We fitted multivariable linear regressions to determine whether study arm was associated with depression or anxiety symptom score change. Covariates included in the regressions included age, genetic sex, genetic ancestry, income estimated at the level of ZCTA, baseline depression symptom score, baseline anxiety symptom score, prior diagnosis of depression, and prior diagnosis of anxiety. To account for heteroskedasticity, we used robust standard errors via the HC3 estimator.(31,32)

Following regression, we implemented the two one-sided tests (TOST) procedure to test for equivalence, in order to provide evidence against the null hypothesis that the study arms were not equivalent in the change in symptoms from baseline to follow-up.(33–36) Specifically, we tested whether the lower bound of the 90% confidence interval for the adjusted treatment effect estimate was greater than the lower bound of the equivalence margin, and whether the upper bound of the 90% confidence interval was less than the upper bound of the equivalence margin. The equivalence bounds were defined as the standard deviation of score changes SD(change)*0.5, based on Cohen’s d = ±0.5, which equates to 1.89 points on the PHQ-8 and 1.13 points on the DASS-21.(34,37–39) Cohen’s d = ±0.5 is generally considered a moderate effective size and is comparable to the equivalence margins defined for prior studies using an equivalence testing framework in psychological research.(34,37–39) Given the SD(change) differed between the exposed and unexposed arms (**Table 1**), we used the smaller standard deviation to define the equivalence bounds. Power analyses confirmed that the study was adequately powered to detect changes in depression and anxiety symptom scores that exceeded the prespecified equivalence margin (depression: power > 0.99; anxiety: power > 0.98). In addition, we conducted a post-hoc exploration to identify the smallest effect size at which we had sufficient (>80%) statistical power (**eTable 1**). Both one-sided tests were conducted using a significance level of α = 0.05, corresponding to a 90% confidence interval for the effect of viewing (vs. not viewing) an at-risk depression PRS result on depression and anxiety symptom scores.

**Table 1.** Descriptive characteristics of study participants at baseline, by exposure to depression PRS results.

|  | Exposed<br>(n = 361) | Unexposed<br>(n = 556) |
| --- | --- | --- |
| **Age** | 53.9 (16.0) | 59.9 (17.0) |
| **Genetic sex (female)** | 297 (82.3%) | 396 (71.2%) |
| **Household income of**<br>**zipcode tabulation area** | 62900 (23100) | 64700 (24400) |
| **Genetic ancestry** |  |  |
| European | 331 (91.7%) | 519 (93.3%) |
| Hispanic/Latino | 22 (6.1%) | 27 (4.9%) |
| African American,<br>Asian, or other | 8 (2.2%) | 10 (1.8%) |
| **Prior diagnosis of**<br>**depression** | 229 (63.4%) | 253 (45.5%) |
| **Prior diagnosis of anxiety** | 230 (63.7%) | 253 (45.5%) |
| **Baseline depression**<br>**symptom score (PHQ-8)** |  |  |
| Mean (SD) | 8.0 (6.5) | 5.4 (5.4) |
| Median [IQR] | 6 [9] | 4 [7] |
| **Baseline anxiety**<br>**symptom score (DASS-21**<br>**anxiety subscale)** |  |  |
| Mean (SD) | 3.8 (3.8) | 2.5 (3.2) |
| Median [IQR] | 3 [5] | 1 [3] |

*Unless otherwise specified, continuous variables are reported as mean ± standard deviation; categorical variables as number (percentage). Individuals with missing data were excluded from the final analysis sample.*

All statistical analyses were conducted using R Version 4.0, and the primary tests used the sandwich and lmtest packages.(32,40–42)

## Results

The final analytic sample included 917 individuals who met eligibility criteria, with 361 individuals in the exposed arm (who viewed the depression PRS result) and 556 in the unexposed arm (who did not view the result). Baseline participant characteristics for the final sample are shown in **Table 1**.

The exposed-arm participants were younger (mean [SD], 53.9 [16.0] years vs. 59.9 [17.0] years in the unexposed arm), more likely to be female (82.3% vs. 71.2% in the unexposed arm), and had higher baseline depression and anxiety symptom scores (mean [SD], PHQ-8: 7.96 [6.52] vs. 5.43 [5.44] in the unexposed arm; DASS-21: 3.84 [3.83] vs. 2.45 [3.18] in the unexposed arm). As expected based on the selection of participants by high PRS for depression, there was a high prevalence of self-reported prior diagnosis of depression and anxiety. The prevalences of prior diagnoses of depression and anxiety were higher in the exposed arm (depression: 63.4% vs. 45.5% in the unexposed arm; anxiety: 63.7% vs. 45.5% in the unexposed arm). Although the exposed arm consisted of research participants at a higher-cost service tier with access to the Depression PRS report, the mean ZCTA-level household income was similar between the two arms. The majority of participants had European genetic ancestry (91.7% in the exposed arm and 93.3% in the unexposed arm). The mean time between completion of the two surveys was 28.0 (SD 4.4) weeks (exposed) and 27.6 (SD 4.4) weeks (unexposed).

Score changes from baseline to follow-up were statistically equivalent for individuals exposed to the depression PRS result compared to individuals who were not (**Figure 3**). For changes in depression symptom scores, the standardized covariate-adjusted effect of viewing an at-risk depression PRS result, compared to not viewing, was −0.044 standard deviations (90%CI, −0.15–0.061; unstandardized values [90% CI]: −0.17 [−0.59–0.24] points of PHQ-8 change). (**Figure 3A**). For changes in anxiety symptom scores, the standardized covariate-adjusted effect of viewing an at-risk depression PRS result, compared to not viewing the PRS result, was −0.038 standard deviations (90%CI, −0.14–0.068; unstandardized values [90% CI]: −0.092 [−0.35–0.17] points of DASS-21 change). For both outcomes, both one-sided tests had p < 0.001, indicating that the 90% confidence interval lay within the equivalence bounds of Cohen’s d = ±0.5 (**Figure 3B**). Complete regression output is shown in **eTable 2**.

**Figure 3.**
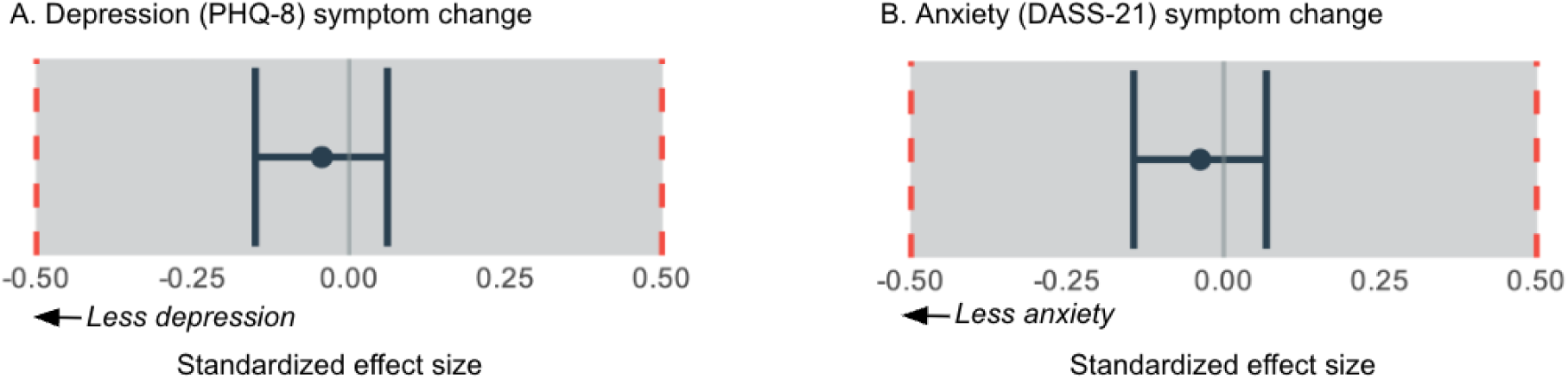
Effects of viewing the PRS on change in depression and anxiety scores. Estimated covariate-adjusted effect of viewing an at-risk depression PRS result versus not viewing on (A) depression symptom change and (B) anxiety symptom change from baseline to follow-up. Dots indicate mean, whiskers indicate the 90% confidence interval, and red dashed lines indicate the prespecified equivalence bounds.

Findings were robust to more elaborate statistical modelling that included additional covariates: time from baseline to follow-up survey completion, an interaction term between viewing the PRS result and time from baseline to follow-up, and interaction terms between viewing the PRS result and prior diagnosis of depression or anxiety (**eTable 3**).

To explore whether viewing the depression PRS result could have a time-varying impact on depression or anxiety symptom scores, such as an initial increase in symptoms that declined over time, we fitted a regression to model the impact of time from exposure to follow-up on score changes among individuals who viewed the PRS result (**eFigure 1**), in addition to all covariates from the primary analysis model. The median time to follow-up survey completion after viewing the depression PRS result was 13.7 weeks (range: 2.3–24.5 weeks). This analysis did not provide evidence of an association between time from exposure to follow-up and changes in depression or anxiety (both p > 0.2).

Finally, we repeated the primary analysis restricting the sample to those who reported a prior diagnosis of depression or anxiety (N = 570) or to individuals who did not (N = 347) (**Figure 4**). In strata defined by presence or absence of previously-diagnosed anxiety or depression, findings were generally consistent with each other (p-het = 0.34 and 0.19 for PHQ-8 and DASS-21, respectively).

**Figure 4.**
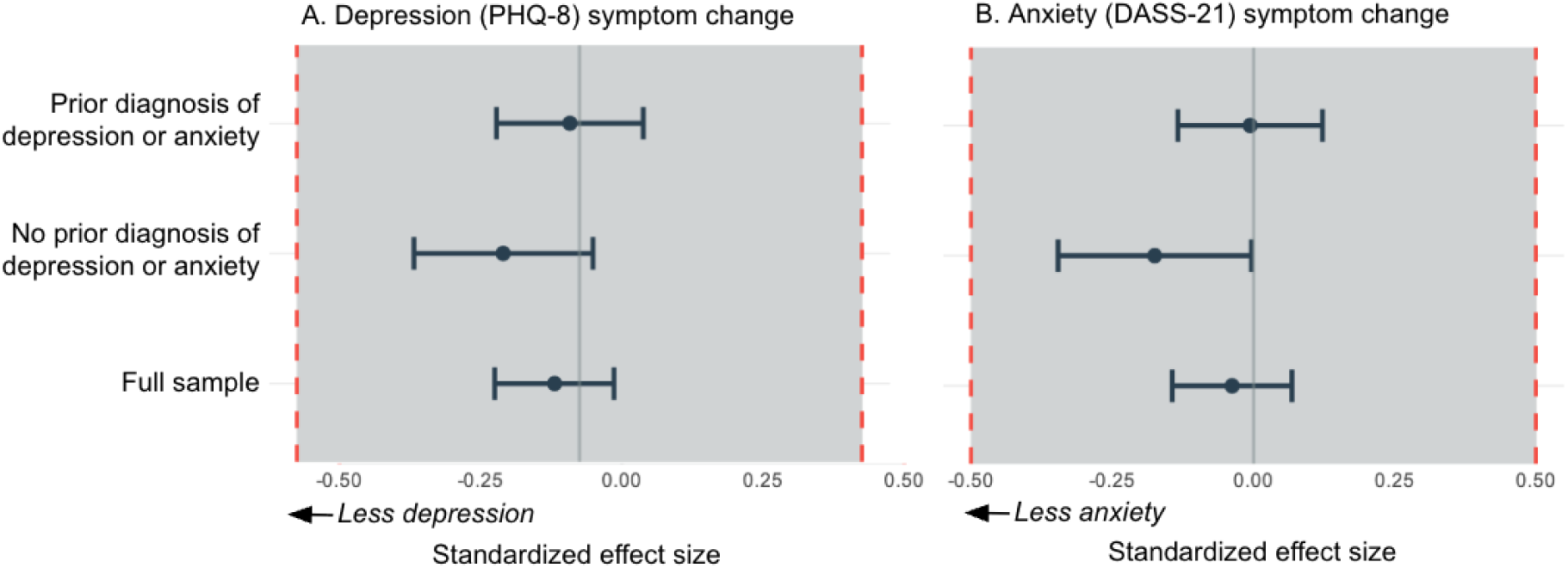
Effects of viewing the PRS on change in depression and anxiety scores in individuals with vs. without a prior diagnosis of depression or anxiety. Estimated covariate-adjusted effect of viewing an at-risk depression PRS result versus not viewing, for subgroups of study participants with (top) or without (middle) a prior diagnosis of depression and/or anxiety, compared the full study sample (bottom). A, estimated effect on PHQ-8 depression symptom scores. B, estimated effect on DASS-21 anxiety symptom scores. Dots represent the mean, whiskers indicate the 90% confidence interval, and red dashed lines indicate the prespecified equivalence bounds.

## Discussion

Depression, a leading cause of disability worldwide,(43) is a frequent focus of consumer interest in genetic testing.(24) It has long been posited that learning of genetic risks, including for psychiatric conditions, may cause psychological distress.(44–46) Two recent surveys found that most psychiatric clinicians were concerned or uncertain about potential psychosocial harms of at-risk PRS results.(46,47) However, consumers consistently report high interest in receiving personal genetic health risk information, including psychiatric genetic information, often seeking to understand current symptoms or family history.(24,48–50) In this large, longitudinal study, we found no sustained impact on depression or anxiety symptoms after receiving a DTC depression PRS result. These findings support the psychological tolerability of delivering this type of information in real-world settings.

Psychological responses to health-related genetic information are shaped by factors such as prior beliefs(51) and how results are framed.(14,52) Although psychiatric conditions are multifactorial, some individuals may interpret genetic risk information deterministically, reducing perceived agency to manage symptoms.(14,21,53) One study found that a brief educational intervention eliminated pessimistic responses to sham depression PRS results.(21) In our study, although results were not accompanied by clinical counseling, participants received contextualizing information—including icon arrays representing absolute risk estimates,(52,54) explanation of depression’s multifactorial etiology, and suggestions for behaviors that can help manage depression symptoms (**Supplement**), which may have helped mitigate negative effects. Future research should explore how psychiatric PRS affect beliefs about genetics and self-efficacy, and identify best practices for presenting results in DTC settings.

This study has several limitations. First, participants were not randomized to study arms, and demographic differences existed across study arms. We adjusted for key covariates, but residual confounding remains possible. Second, we assessed symptoms 6 months post-disclosure, so short-term emotional responses could have been missed. Prior studies have found brief increases in test-specific anxiety following genetic risk disclosures,(55) but we observed no association between time since result receipt and symptom change. Finally, our cohort was disproportionately white and of high socioeconomic status, and those who completed follow-up surveys may differ systematically from the broader population, raising the possibility of selection bias.

This study also has several strengths. First, we used an equivalence testing framework to rigorously evaluate whether receiving a PRS result was associated with clinically meaningful changes in symptoms. This method tests whether outcomes fall within a prespecified equivalence margin—in this case, Cohen’s d = ±0.5, which corresponds to <2 points on the PHQ-8 and DASS-21 and likely falls below the threshold for clinical significance.(34,37–39) Our findings were well within this range, and post-hoc power calculations showed sufficient power rule out even smaller effects (Cohen’s d = ±0.3; **eTable 1**).

Second, our survey design minimized response bias by separating symptom measurement from the PRS result experience. Surveys were administered via a different webpage and login session, with no content explicitly linking the depression PRS result to the symptom questions. This independent presentation reduces the likelihood that participants’ responses were primed by result-related expectations.

Third, this is one of the first large-scale studies to examine psychiatric PRS disclosure in a real-world, population-based setting. Unlike clinically selected populations receiving genetic testing for a specific condition, our participants were not necessarily seeking genetic risk information for depression. At the time of report release, they had access to >140 previously released reports across health conditions, pharmacogenetics, traits, and ancestry. In this context, compared to clinical testing, individuals may be less prepared to receive an at-risk result and could have less access to clinical support, which could plausibly increase psychological vulnerability. Despite this, we found no evidence of adverse psychological effects.

Our findings align with past research showing minimal psychosocial impacts from genetic testing for cardiovascular, neurodegenerative, and cancer conditions in clinical settings.(10,11,51,55–57) Our study extends this literature by addressing psychiatric PRS in a real-world setting.

Future research should assess whether these findings generalize to more racially, socioeconomically, and clinically diverse populations, including individuals with more severe psychiatric symptoms or strong family histories. While our study focused on potential harms, future research should also evaluate potential benefits. Psychiatric PRS may hold promise for supporting risk assessment, improving diagnosis, guiding preventive strategies, and predicting treatment response,(58–60) but robust evidence of clinical utility in diverse populations is still needed.

In conclusion, we found no evidence of clinically meaningful changes in depression or anxiety symptoms following receipt of psychiatric PRS in a large, real-world, DTC setting. These findings provide important information about the psychological safety of returning psychiatric genetic risk information outside of a clinical context, and suggest a foundation for exploring future applications of mental health genetic risk assessments in healthcare.

## Supporting information

Supplementary Content

## Data Availability

Individual-level data are not publicly available, due to participant privacy, and in accordance with the IRB-approved protocol under which the study was conducted.

## Acknowledgements

We thank the research participants of 23andMe for making this work possible. The following members of the 23andMe Genomic Health and Sciences, Product R&D, and Research teams contributed to this study: James Ashenhurst, Stella Aslibekyan, Adam Auton, Robert Bell, Jessica Bielenberg, Kayla Capper, Zayn Cochinwala, Kahsaia de Brito, Emily DelloRusso, Stacey Detweiler, Sarah Elson, Shirin Fuller, Julie M. Granka, Barry Hicks, David Hinds, Lucy Kaufmann, Bertram Koelsch, Sarah Laskey, Alisa Lehman, Hannah Llorin, Matthew McIntyre, Carrie Northover, Marlena Ortlieb, Jamaica Perry, Katie Sagaser, Anjali Shastri, Teague Sterling, Joyce Tung, Vinh Tran, Shirley Wu, Jianan Zhan. We thank Rafaela Bagur Quetglas for developing the depression polygenic risk score model.

## Declaration of generative AI and AI-assisted technologies in the writing process

During the preparation of this work the authors used ChatGPT to provide minor editing suggestions to improve the readability of the text. After using this tool, the authors reviewed and edited the content as needed and take full responsibility for the content of the publication.

