## Supplementary Content for "Viewing Direct-to-Consumer Genetic Test Results for Depression Risk Is Psychologically Well Tolerated: Evidence from a Longitudinal Equivalence Study"

**eTable 1.** Post-hoc power analysis conducted at a range of effect sizes.

**eTable 2.** Linear regression of changes in depression and anxiety symptom scores by exposure to at-risk depression PRS results

**eTable 3.** Linear regression of changes in depression and anxiety symptom scores by exposure to at-risk depression PRS results, with additional covariates

**eFigure1.** Change in depression and anxiety symptom score by time between at-risk depression PRS result viewing and follow-up survey

**eTable 1. Post-hoc power analysis conducted at a range of effect sizes, for the final study sample.**

| **Outcome** | **Cohen's d** | **Effect size in raw points** | **Power** |
| --- | --- | --- | --- |
| Depression score change (PHQ-8) | 0.2 | 0.75 | 0.38 |
|  | 0.3 | 1.13 | 0.88 |
|  | 0.4 | 1.51 | >0.99 |
|  | 0.5 | 1.89 | >0.99 |
| Anxiety score change (DASS-21) | 0.2 | 0.45 | 0.48 |
|  | 0.3 | 0.67 | 0.92 |
|  | 0.4 | 0.90 | >0.99 |
|  | 0.5 | 1.13 | >0.99 |

**eTable 2. Linear regression of changes in depression and anxiety symptom scores by exposure to at-risk depression PRS results**

|  | **Depression score change** | | | **Anxiety score change** | | |
| --- | --- | --- | --- | --- | --- | --- |
|  | **β** | **SE** | **p** | **β** | **SE** | **p** |
| **Age** | -0.025 | 0.0089 | 0.004** | -0.018 | 0.005 | <0.001*** |
| **Genetic sex (male)** | -0.16 | 0.29 | 0.57 | -0.24 | 0.18 | 0.09 |
| **Household income of zipcode tabulation area** | -0.00001 | 0.000004 | 0.01* | -0.000006 | 0.000003 | 0.04* |
| **Genetic ancestry group** |  |  |  |  |  |  |
| **Hispanic/Latino** | 0.19 | 0.56 | 0.73 | 0.64 | 0.33 | 0.05 |
| **African American, Asian, or other** | 0.057 | 0.46 | 0.90 | -0.002 | 0.53 | 0.10 |
| **Prior diagnosis of depression** | 1.35 | 0.29 | <0.001*** | 0.23 | 0.19 | 0.19 |
| **Prior diagnosis of anxiety** | 0.47 | 0.30 | 0.12 | 0.43 | 0.20 | 0.02* |
| **Baseline depression symptom score (PHQ-8)** | -0.37 | 0.037 | <0.001*** | 0.095 | 0.018 | <0.001*** |
| **Baseline anxiety symptom score (DASS-21 anxiety subscale)** | 0.13 | 0.067 | 0.05 | -0.045 | 0.031 | <0.001*** |
| **Exposure to depression PRS result** | -0.17 | 0.25 | 0.49 | -0.092 | 0.15 | 0.56 |

*Standard errors and p-values are computed using the sandwich method to adjust for heteroskedasticity. p < .05 = *, p < .01 = **, p < .001 = ****

**eTable 3. Linear regression of changes in depression and anxiety symptom scores by exposure to at-risk depression PRS results, with additional covariates**

|  | **Depression score change** | | | **Anxiety score change** | | |
| --- | --- | --- | --- | --- | --- | --- |
|  | **β** | **SE** | **p** | **β** | **SE** | **p** |
| **Age** | -0.026 | 0.0090 | 0.003** | -0.018 | 0.0054 | <0.001*** |
| **Genetic sex (male)** | -0.18 | 0.29 | 0.54 | -0.24 | 0.14 | 0.09 |
| **Household income of zipcode tabulation area** | -0.000012 | 0.0000049 | 0.01* | -0.0000060 | 0.0000028 | 0.03* |
| **Genetic ancestry group** |  |  |  |  |  |  |
| **Hispanic/Latino** | 0.20 | 0.57 | 0.72 | 0.62 | 0.33 | 0.06 |
| **African American, Asian, or other** | -0.0061 | 0.47 | 0.99 | -0.012 | 0.42 | 0.98 |
| **Prior diagnosis of depression** | 1.3 | 0.39 | <0.001*** | 0.24 | 0.21 | 0.27 |
| **Prior diagnosis of anxiety** | 0.41 | 0.40 | 0.30 | 0.29 | 0.22 | 0.19 |
| **Baseline depression symptom score (PHQ-8)** | -0.37 | 0.038 | <0.001*** | 0.094 | 0.023 | <0.001*** |
| **Baseline anxiety symptom score (DASS-21 anxiety subscale)** | 0.13 | 0.068 | 0.05 | -0.45 | 0.049 | <0.001*** |
| **Exposure to depression PRS result** | -3.2 | 1.6 | 0.05 | -0.77 | 1.1 | 0.47 |
| **Time between survey instances** | -0.0053 | 0.0058 | 0.36 | 0.0018 | 0.0037 | 0.64 |
| **Exposure to depression PRS result : prior diagnosis of depression [interaction]** | 0.19 | 0.56 | 0.74 | 0.014 | 0.36 | 0.97 |
| **Exposure to depression PRS result : prior diagnosis of anxiety [interaction]** | 0.14 | 0.56 | 0.81 | 0.35 | 0.36 | 0.32 |
| **Exposure to depression PRS result : time between survey instances [interaction]** | 0.014 | 0.0084 | 0.09 | 0.0024 | 0.0054 | 0.66 |

*Standard errors and p-values are computed using the sandwich method to adjust for heteroskedasticity. p < .05 = *, p < .01 = **, p < .001 = ****

**eFigure 1. Change in depression and anxiety symptom scores by time between at-risk depression PRS result viewing and follow-up survey**

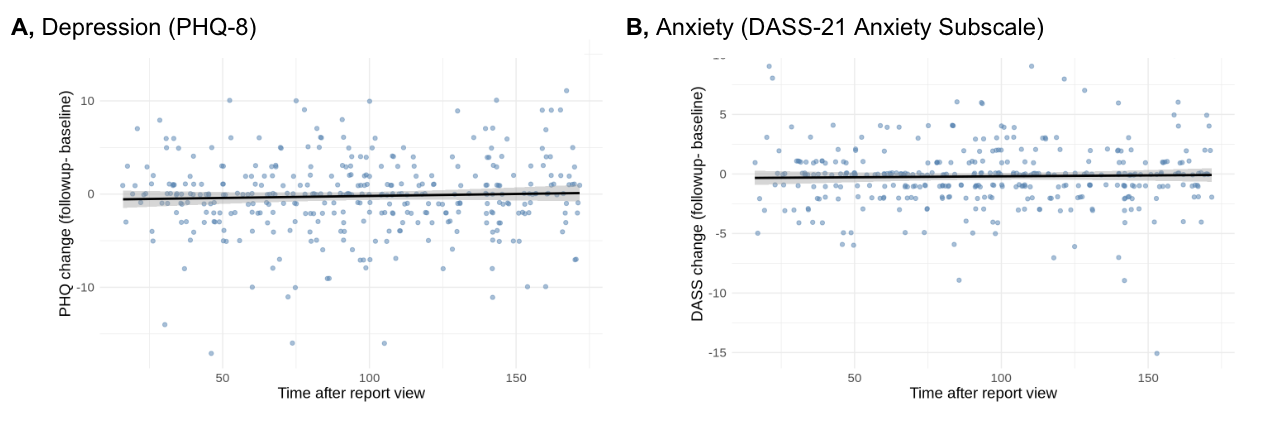

Change in (A) depression and (B) anxiety symptoms by time since viewing at-risk depression PRS report, among individuals who viewed the depression PRS report. Solid line shows the unadjusted fitted linear regression; shaded area indicates the 95% confidence interval for the predicted mean change in PHQ-8 or DASS-21 scores, widening with distance from the mean of the time variable due to increasing uncertainty.
